# SARS-CoV-2 seropositivity and seroconversion in patients undergoing active cancer-directed therapy

**DOI:** 10.1101/2021.01.15.21249810

**Authors:** Lova Sun, Sanjna Surya, Noah G. Goodman, Anh N. Le, Gregory Kelly, Olutosin Owoyemi, Heena Desai, Cathy Zheng, Shannon DeLuca, Madeline L. Good, Jasmin Hussain, Seth D. Jeffries, Yolanda R. Kry, Emily M. Kugler, Maikel Mansour, John Ndicu, AnnaClaire Osei-Akoto, Timothy Prior, Stacy L. Pundock, Lisa A. Varughese, JoEllen Weaver, Abigail Doucette, Scott Dudek, Shefali Setia Verma, Sigrid Gouma, Madison E. Weirick, Christopher M. McAllister, Erin Bange, Peter Gabriel, Marylyn Ritchie, Daniel J. Rader, Robert H. Vonderheide, Lynn M Schuchter, Anurag Verma, Ivan Maillard, Ronac Mamtani, Scott E. Hensley, Robert Gross, E. Paul Wileyto, Alexander C. Huang, Kara N. Maxwell, Angela DeMichele

## Abstract

Multiple studies have demonstrated the negative impact of cancer care delays during the COVID-19 pandemic, and transmission mitigation techniques are imperative for continued cancer care delivery. To gauge the effectiveness of these measures at the University of Pennsylvania, we conducted a longitudinal study of SARS-CoV-2 antibody seropositivity and seroconversion in patients presenting to infusion centers for cancer-directed therapy between 5/21/2020 and 10/8/2020. Participants completed questionnaires and had up to five serial blood collections. Of 124 enrolled patients, only two (1.6%) had detectable SARS-CoV-2 antibodies on initial blood draw, and no initially seronegative patients developed newly detectable antibodies on subsequent blood draw(s), corresponding to a seroconversion rate of 0% (95%CI 0.0-4.1%) over 14.8 person-years of follow up, with a median of 13 healthcare visits per patient. These results suggest that cancer patients receiving in-person care at a facility with aggressive mitigation efforts have an extremely low likelihood of COVID-19 infection.

## Introduction

Patients with cancer are at risk for poor outcomes with COVID-19[1-9]. In order to reduce person-to-person contact, a wide array of cancer care procedures have been altered or delayed during the pandemic[10, 11]. However, these disruptions are projected to lead to almost 10,000 excess deaths over the next decade from breast and colorectal cancer alone[12, 13] – a grim second public health crisis arising as a consequence of the pandemic. Patients with cancer face difficult decisions between continuing cancer-directed care and avoiding healthcare settings to decrease infection risk.

The clear need to continue delivering cancer care while minimizing risk of SARS-CoV-2 infection has highlighted the importance of transmission mitigation techniques in both healthcare and community settings. At outpatient oncology clinics at the University of Pennsylvania (Penn), efforts to reduce in-person patient volume, including virtual visits, at-home infusions, and decreased infusion frequencies, began in mid-March, 2020. For oncology patients who continued to receive in-person care, safety measures included text message questionnaires regarding contact with COVID-positive individuals, temperature screenings, contactless check-in procedures, visitor limitations, and physically separated COVID-19+ clinic spaces.

## Methods

In order to gauge the effectiveness of these measures, we conducted a prospective longitudinal study to assess the rate of SARS-CoV-2 antibody seropositivity and seroconversion in patients undergoing active cancer therapy. Patients with solid malignancies presenting to the Penn Abramson Cancer Center for chemotherapy and/or immunotherapy were enrolled between 5/21/2020 and 10/8/2020. Patients were approached via phone and provided electronic consent for blood collection, electronic surveys, and access to electronic health records (EHR). We targeted patients receiving therapy on a single day of the week to capture a mixture of solid malignancy types and facilitate longitudinal follow-up.

In order to assess seropositivity and seroconversion, up to five serial blood draws were performed during treatment visits at a minimum of three weeks apart. At each encounter, 50mL of blood was drawn and centrifuged for 15 minutes; serum, plasma and buffy coat were stored at −80C. Samples were assayed for SARS-CoV-2 IgG and IgM antibodies to the spike receptor binding domain antigen using an enzyme-linked immunosorbent assay (ELISA) approach, with high sensitivity and specificity[14]. Patients received electronic questionnaires at each of their blood draw dates which assessed self-reported measures of health, SARS-COV-2 exposure, and social interactions. Patients were not informed of antibody test results. This study was approved by the institutional review board at Penn.

## Results

Of 440 eligible patients approached, 124 (28%) consented and had antibody testing. The remainder either could not be reached (n=89, 28%) or declined participation (n=227, 72%). Most enrolled patients had metastatic cancer, with a median of two prior lines of systemic therapy; approximately half had ECOG performance status>0; and over a third were leukopenic over the course of the study **(Table 1)**. Patients had a median of 13 (IQR 9-18) in-person healthcare visits and seven (IQR 5-10) cycles of cancer therapy from March to final study blood draw, based on EHR review. Of patients who completed the initial questionnaire, 110/121 (94%) reported being highly compliant (4-5 on Likert scale) with CDC-recommended social distancing measures, and 81/118 (69%) reported a low level of social interactions (going out ≤3 times a week) **(Supplemental Table 1)**. Seventy-eight patients completed ≥1 subsequent survey; the majority maintained low level of social interactions (57%) or decreased their interactions to ≤3 times per week (19%) (**Supplemental Table 2**). Most patients reported no exposures to individuals known or suspected to have SARS-CoV-2 infection on initial (n=102, 85%) and subsequent surveys (n=66, 85%).

**Table 1.** Study Subjects

| **Characteristic** | **Number (%), N=124** |
| --- | --- |
| Age, years (IQR) | 62 (53-69) |
| Female Sex | 64 (52%) |
| Race |  |
| White | 104 (84%) |
| Black or African American | 10 (8%) |
| Asian | 4 (3%) |
| Multiple Races | 1 (1%) |
| Other/Unknown | 5 (4%) |
| Hispanic Ethnicity | 2 (2%) |
| ECOG performance status |  |
| 0 | 63 (51%) |
| 1 | 44 (35%) |
| 2 | 10 (8%) |
| 3 | 7 (6%) |
| Smoking history |  |
| Never | 61 (49%) |
| Prior | 54 (44%) |
| Current | 9 (7%) |
| Charlson Comorbidity Index, median (IQR) | 8 (6-9) |
| BMI, median (IQR) | 27 (23-32) |
| Primary Cancer Diagnosis |  |
| Lung | 28 (23%) |
| Breast | 22 (18%) |
| Melanoma | 19 (15%) |
| Kidney | 17 (14%) |
| Colon/Rectum | 13 (11%) |
| Pancreas | 10 (8%) |
| Prostate | 4 (3%) |
| Esophagus or Gastric | 4 (3%) |
| Other<sup>a</sup> | 7 (6%) |
| Stage of cancer |  |
| I | 9 (7%) |
| II | 7 (6%) |
| III | 35 (28%) |
| IV | 71 (57%) |
| Unknown | 2 (2%) |
| Number of lines of systemic therapy, median (IQR) | 2 (1-4) |
| Cancer Therapy Type |  |
| Chemotherapy | 62 (50%) |
| Immunotherapy | 54 (44%) |
| Chemoimmunotherapy | 8 (7%) |
| Number of visits since March 1, median (IQR) | 13 (9-18) |
| Number of therapy cycles since March 1, median (IQR) | 7 (5-10) |
| Neutropenic (ANC< 500) since March 1 | 3 (2%) |
| Leukopenic (WBC< LLN) since March 1 | 46 (37%) |
| Number of blood draws for SARS-CoV-2 antibodies |  |
| 2 | 90 (73%) |
| 3 | 51 (41%) |
| 4 | 21 (17%) |
| 5 | 8 (6%) |

<sup>a</sup>Other cancers included ovary (n=1), pharynx (n=1), thymus (n=1), and unknown primary (n=4).
Abbreviations: IQR, interquartile range; ECOG, Eastern Cooperative Oncology Group; BMI, body mass index; ANC, absolute neutrophil count; WBC, white blood cell; LLN, lower limit of normal

Of 124 patients enrolled, 90 had at least one subsequent blood draw, a median of 28 days apart, comprising 14.9 person-years of follow up **(Figure 1)**. Only two (1.6%) participants had detectable antibodies to SARS-CoV-2 on their first blood draw. Both of these patients were female, white, and receiving chemotherapy for metastatic cancer. One patient with breast cancer had symptomatic COVID-19 in April and had only detectable IgG on her two on-study blood draws (8/27 and 9/24). The other patient, with esophageal cancer, had only detectable IgM on her two blood draws (8/20 and 9/17) and never developed symptomatic COVID-19. Of 88 seronegative patients with at least one subsequent blood draw, none developed detectable antibodies to SARS-CoV-2 on any subsequent blood draw, corresponding to a seroconversion rate of 0% (95% CI 0-4.1%) over 14.8 person-years of follow up.

**Figure 1.**
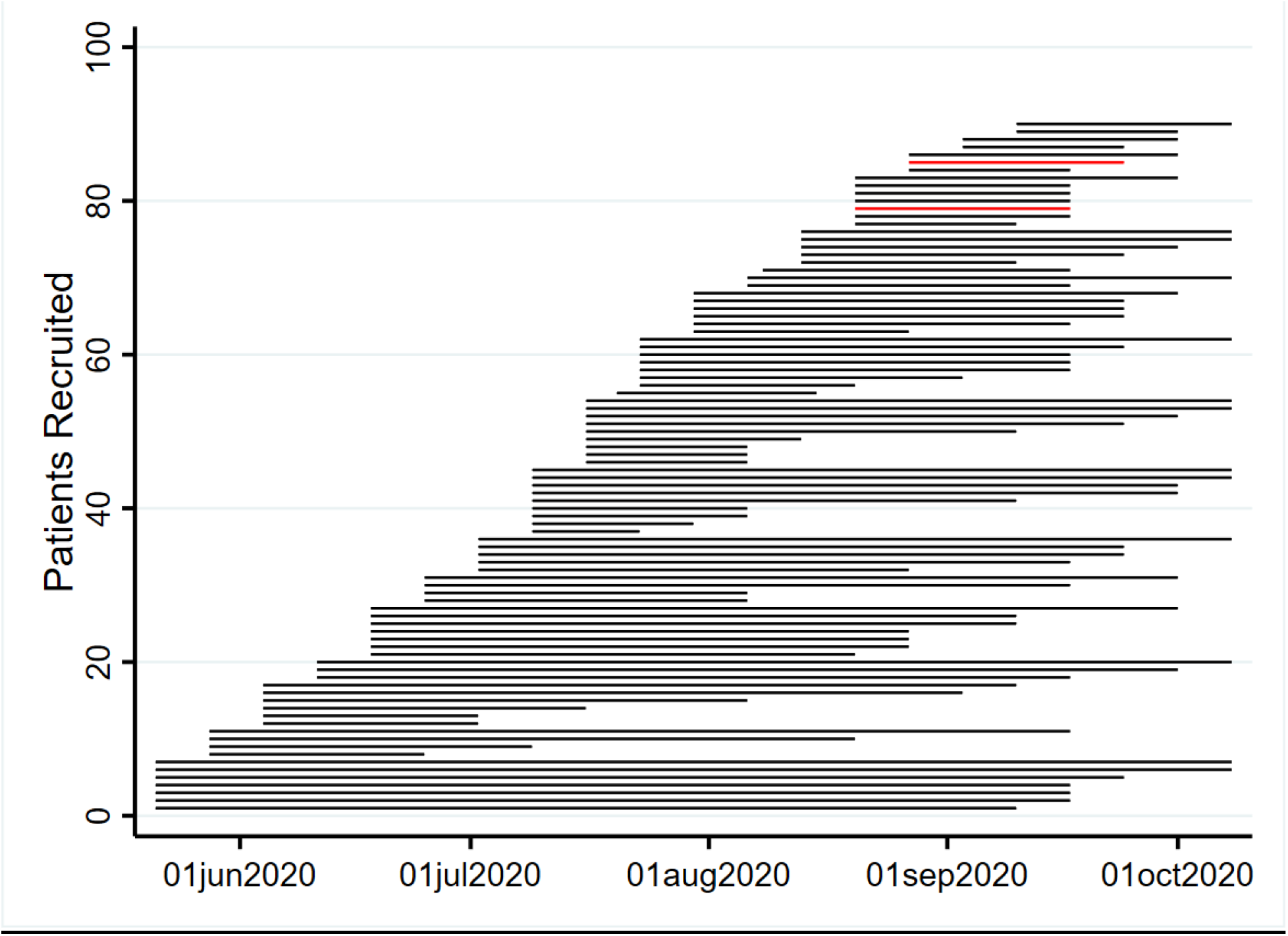
Swimmers Plot: Time under observation for seroconversion in patients with at least one serial blood draw (n=90): Each patient’s time under observation is represented by a horizontal bar, which spans the time from first to final blood draw. Black represents seronegative; red represents seropositive (detectable anti-SARS-CoV-2 IgG or IgM antibody). No patients who were initially seronegative became seropositive on subsequent blood draw.

## Discussion and Conclusion

In this single-center prospective study, we found a low rate of seropositivity and no seroconversions in patients interacting frequently with the healthcare system in-person to receive active cancer-directed therapy. Other reports of SARS-CoV-2 seroprevalence in cancer patients have ranged from 3.6%[15] to 31.4%[16]. These reports dated from periods and locations when population estimates were higher than that in Philadelphia during the study period; the non-cancer US population had an estimated seroprevalence <10% in the majority of jurisdictions studied from August-September 2020[17]. However, a study of parturient women giving birth at two Philadelphia hospitals from April-June 2020 reported a higher 6.2% seroprevalence rate[14].

Serologic assays are imperfect measures of SARS-CoV-2 exposure and infection due to variable test sensitivity, timing, and antibody persistence[18, 19]. In particular, patients with less severe illness, as well as cancer patients on cytotoxic or immunomodulatory therapy, may have lower SARS-CoV-2 antibody levels and detection rates[20, 21]. Moreover, seroprevalence rates are a product of multiple demographic, geographic, temporal, and behavioral factors, and thus it is not possible to determine the precise causal relationship between health system mitigation strategies and seroconversion without a randomized trial. Questionnaire responses showed that our patients were generally highly adherent to COVID-19 prevention strategies, potentially due to a personal sense of vulnerability. However, the low rate of SARS-CoV-2 seropositivity and seroconversion seen in our cohort likely also reflects the success of transmission mitigation measures within healthcare facilities, and suggests that these efforts, when combined with social distancing outside the healthcare setting, may help protect vulnerable cancer patients from SARS-CoV-2 exposure and infection, even when ongoing immunomodulatory cancer treatments and frequent healthcare exposure are necessary. Continued reinforcement of practices including physical distancing, masking, and visitor limitation will remain critical as pandemic fatigue rises along with COVID-19 cases.

## Data Availability

Data available on request. The data underlying this article will be shared on reasonable request to the corresponding author.

## Acknowledgements

We acknowledge the Penn Medicine BioBank (PMBB) for providing data and thank the patient-participants of Penn Medicine who consented to participate in this research program. The PMBB is approved under IRB protocol #813913.

## Author Disclosures

RHV reports having received consulting fees or honoraria from Medimmune and Verastem; and research funding from Fibrogen, Janssen, and Lilly. He is a member of the Lustgarten Therapeutics Advisory working group. He is an inventor on a licensed patents relating to cancer cellular immunotherapy and cancer vaccines, and receives royalties from Children’s Hospital Boston for a licensed research-only monoclonal antibody. SEH has received consultancy fee from Sanofi Pasteur, Lumen, Novavax, and Merck for work unrelated to this report. Other authors declare that they have no competing interests.

## Notes

***Funding:*** This work was supported by the National Institutes of Health (P30-CA016520 to Abramson Cancer Center and RHV), a gift from the Smilow family, the National Center for Advancing Translational Sciences of the National Institutes of Health under CT6SA Award Number UL1TR001878, the Tara Miller Melanoma Excellence Fund, and the Perelman School of Medicine at the University of Pennsylvania. J. Lurie, J. Embiid, J. Harris, and D. Blitzer provided philanthropic support that was critical for establishing the serological assays used in this study.

### Funding Statement

This work was supported by the National Institutes of Health (P30-CA016520 to Abramson Cancer Center and RHV), a gift from the Smilow family, the National Center for Advancing Translational Sciences of the National Institutes of Health under CT6SA Award Number UL1TR001878, the Tara Miller Melanoma Excellence Fund, and the Perelman School of Medicine at the University of Pennsylvania. J. Lurie, J. Embiid, J. Harris, and D. Blitzer provided philanthropic support that was critical for establishing the serological assays used in this study.

### Author Declarations

This study was approved by the institutional review board at Penn Medicine under IRB protocol #813913.

